# Factors Influencing Hospital Nurses Performance In Accordance With Clinical Authority: A Literature Review

**DOI:** 10.1101/2025.09.11.25335258

**Authors:** Menik Kustriyani, Arief Yanto

## Abstract

**Background:** Hospital nurses performance within their clinical authority is pivotal to patient safety and care quality. Recent scholarship (2021-2025) indicates that performance is not solely a function of individual competence but is shaped by a dynamic interplay of organizational, individual, and environmental factors.

**Purpose:** This literature review aimed to synthesize current evidence on the determinants that facilitate or impede hospital nurses performance in accordance with their clinical authority, and to highlight gaps for future research.

**Methods:** A systematic search of peer-reviewed articles published between 2021 and 2025 was conducted across major nursing and health services databases. Inclusion criteria comprised empirical studies (qualitative, quantitative, or mixed methods) that examined factors influencing nurses performance, leadership, empowerment, workload, or environmental considerations. Selected articles were screened, coded, and thematically analyzed to identify recurring determinants, barriers, and facilitators.

**Results:** The synthesis revealed four core determinants: (1) supportive clinical leadership, which exerts the strongest direct and indirect influence on performance (0.35); (2) manageable workloads, mitigating role conflict between patient care priority and environmental stewardship; (3) structural empowerment and professional autonomy, linked to innovative behaviours (0.63) mediated by organizational climate; and (4) positive organizational climate/team climate, fostering collaboration. Prominent barriers included hierarchical cultures, resource shortages, and insufficient mentorship. Environmental awareness, while present, was often subordinated to immediate clinical duties due to time constraints and lack of institutional support.

**Conclusion:** Hospital nurses performance under clinical authority is governed by a complex nexus of leadership, workload, empowerment, and climate factors. Targeted organizational interventions, coupled with rigorous research in diverse contexts, are essential to optimize nursing performance and sustain high-quality patient care.

## Introduction

Hospital nurses’ performance, particularly in exercising their clinical authority, is a cornerstone of healthcare quality and patient safety. Adhering to clinical authority allows nurses to deliver care that matches their training and credentialed competencies, leading to improved patient outcomes and satisfaction. Empowering nurses with appropriate clinical authority is consistently linked to better patient care and more effective health systems, as it enables timely, evidence-based decision-making and interventions (Pujianti et al., 2024; Yanto and Kustriyani, 2023). When nurses operate within their defined scope, patient safety is enhanced, and the risk of errors or adverse events is reduced (Supri et al., 2019).

Clinical authority refers to the scope and autonomy nurses possess to make clinical decisions and perform interventions within their professional competencies. Working within clinical authority fosters professionalism and confidence among nurses. Nurses who practice according to their competencies are more likely to feel confident, satisfied, and motivated, which in turn positively affects their performance and patient satisfaction (Supri et al., 2019). Structured career paths and clear clinical authority also promote responsibility, adherence to standards, and mutual trust within healthcare teams (Pujianti et al., 2024). Understanding the factors that influence nurses’ ability to perform in accordance with their clinical authority is critical, as it directly impacts patient outcomes, staff satisfaction, and organizational effectiveness (Hyun and Lee, 2021; Kalogirou et al., 2021).

The researcher aims to conduct a literature review on the topic “Factors Influencing Hospital Nurses’ Performance in Accordance With Clinical Authority.” The review will synthesize evidence on nine core determinants organizational structure, individual nurse characteristics, work-environment conditions, professional regulation, clinical leadership, advanced education, inter-professional collaboration, motivational aspects, and performance evaluation systems. An additional impetus for this review is that the enactment of clinical authority by nurses in Indonesia remains considerably limited, potentially compromising the quality of nursing care, reducing nurses’ participation in clinical decision-making, and adversely affecting patient safety and outcomes.

## Method

This research uses a literature review approach the databases used are Scopus, Science Direct, PubMed, and Dimensions. The strategy used to search the literature by enter the keywords: nurse* OR nursing OR “registered nurse*” OR RN AND performance* OR outcome* OR effectiveness OR quality OR productivity OR efficiency AND authority* OR empower* OR autonom* OR “decision making” OR leadership OR power.

Inclusion criteria were: articles published in English between 2021 and 2025, full-text availability, and relevance to the themes of nurses performance clinical authority. Exclusion criteria were: publications older than five years, review articles, and studies published in Indonesian. There were 4,868 articles from the Scopus database, 10 articles from Sciencedirect, 89 articles from PubMed, and 10 articles from Dimensions, so the total articles obtained are 4,977 articles. After screening based on inclusion and exclusion criteria, 23 research articles were obtained consisting international articles.

**Table 1.**
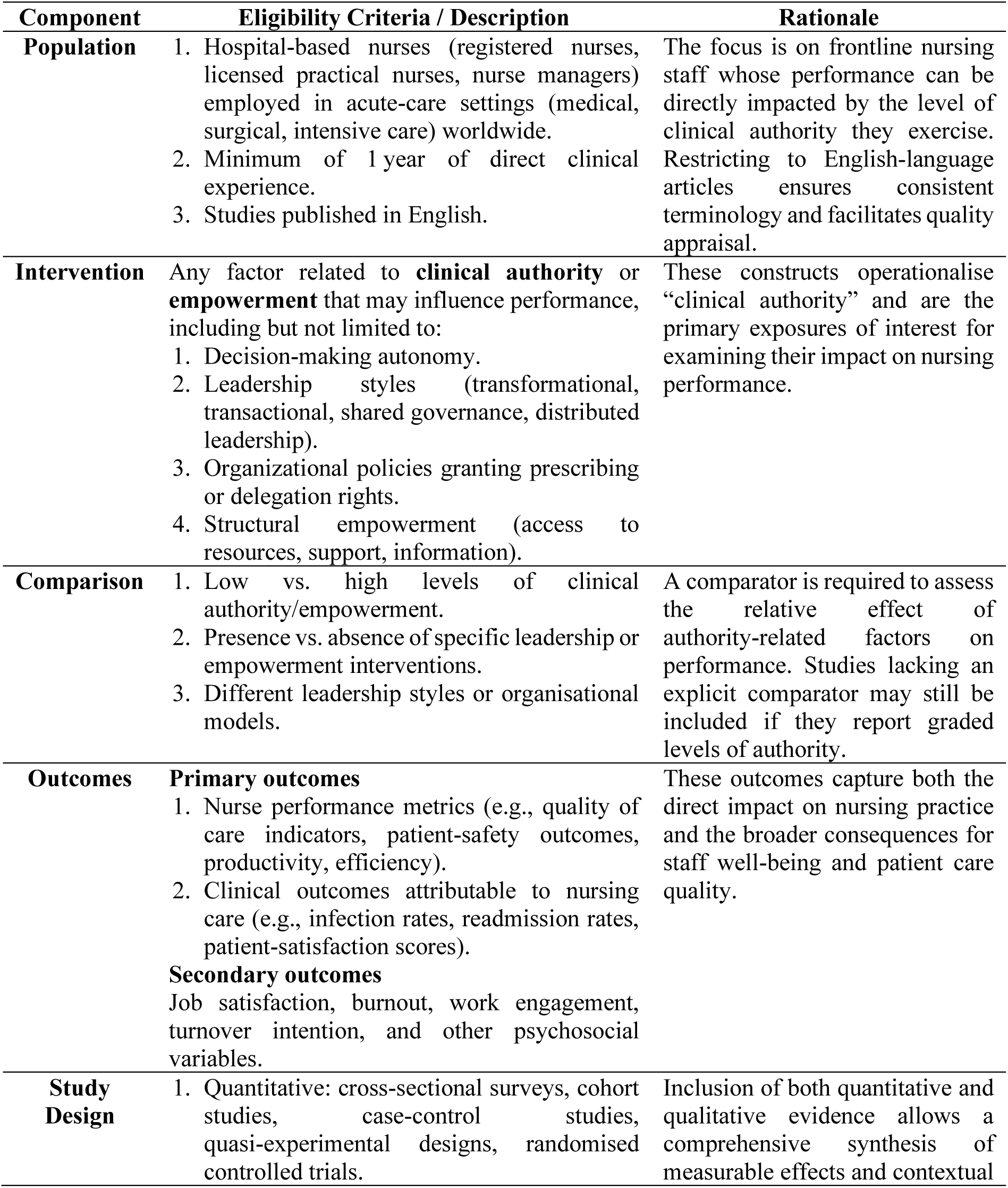
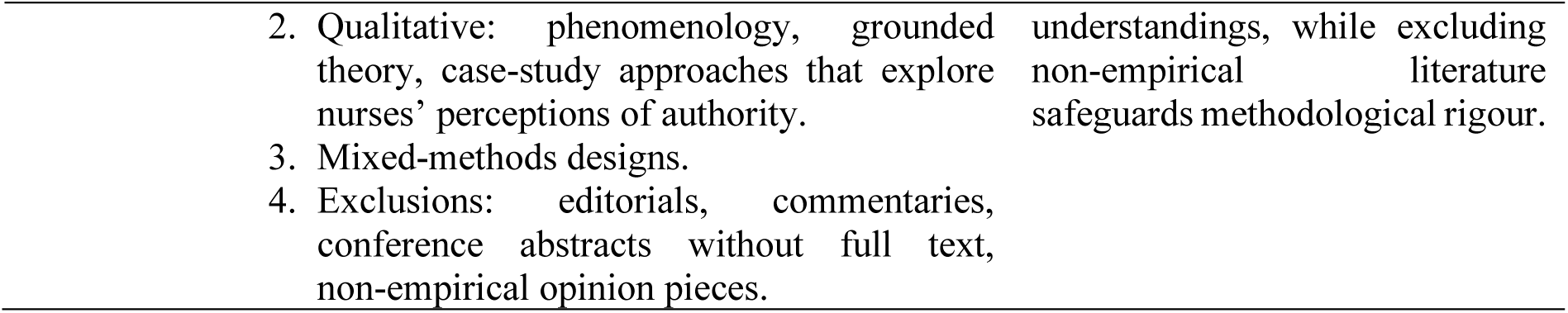
PICOS Table for Article Screening.

| Component | Eligibility Criteria / Description | Rationale |
| --- | --- | --- |
| **Population** | 1. Hospital-based nurses (registered nurses, licensed practical nurses, nurse managers) employed in acute-care settings (medical, surgical, intensive care) worldwide.<br>2. Minimum of 1 year of direct clinical experience.<br>3. Studies published in English. | The focus is on frontline nursing staff whose performance can be directly impacted by the level of clinical authority they exercise.<br>Restricting to English-language articles ensures consistent terminology and facilitates quality appraisal. |
| **Intervention** | Any factor related to **clinical authority** or **empowerment** that may influence performance, including but not limited to:<br>1. Decision-making autonomy.<br>2. Leadership styles (transformational, transactional, shared governance, distributed leadership).<br>3. Organizational policies granting prescribing or delegation rights.<br>4. Structural empowerment (access to resources, support, information). | These constructs operationalise “clinical authority” and are the primary exposures of interest for examining their impact on nursing performance. |
| **Comparison** | 1. Low vs. high levels of clinical authority/empowerment.<br>2. Presence vs. absence of specific leadership or empowerment interventions.<br>3. Different leadership styles or organisational models. | A comparator is required to assess the relative effect of authority-related factors on performance. Studies lacking an explicit comparator may still be included if they report graded levels of authority. |
| **Outcomes** | **Primary outcomes**<br>1. Nurse performance metrics (e.g., quality of care indicators, patient-safety outcomes, productivity, efficiency).<br>2. Clinical outcomes attributable to nursing care (e.g., infection rates, readmission rates, patient-satisfaction scores).<br>**Secondary outcomes**<br>Job satisfaction, burnout, work engagement, turnover intention, and other psychosocial variables. | These outcomes capture both the direct impact on nursing practice and the broader consequences for staff well-being and patient care quality. |
| **Study Design** | 1. Quantitative: cross-sectional surveys, cohort studies, case-control studies, quasi-experimental designs, randomised controlled trials. | Inclusion of both quantitative and qualitative evidence allows a comprehensive synthesis of measurable effects and contextual |
|  | 2. Qualitative: phenomenology, grounded theory, case-study approaches that explore nurses' perceptions of authority. | understandings, while excluding non-empirical literature safeguards methodological rigour. |
|  | 3. Mixed-methods designs. |  |
|  | 4. Exclusions: editorials, commentaries, conference abstracts without full text, non-empirical opinion pieces. |  |

## Results

### 1. Organizational Context

Patient-care priority vs. environmental stewardship: Kalogirou et al. revealed that nurses view patient outcomes as the primary duty, relegating environmental considerations to a secondary concern when workload is high. Implications for performance: When organisational policies do not integrate sustainability into routine workflows, nurses experience role conflict, potentially diminishing overall performance quality.

### 2. Clinical Leadership, Team Climate & Structural Empowerment

Leadership’s central role: Kuşcu Karatepe & Türkmen demonstrated that clinical leadership exerts the strongest standardized influence (β =.35) on performance, both directly and via a supportive creative team climate (β =.23) and structural empowerment (β =.19). Variance explained: Together these factors account for 39 % of performance variability, underscoring leadership as a lever for improvement.

### 3. Empowerment, Autonomy & Innovation

Head-nurse empowerment: Lu et al. found a robust positive relationship (β = 0.635) between head-nurse empowerment and nurses’ innovative behaviour, mediated heavily (≈ 71 %) by organizational climate and professional autonomy. Practical takeaway: Empowering senior nurses and fostering autonomy creates a fertile ground for innovative practice, which in turn enhances performance aligned with clinical authority.

### 4. Environmental Awareness & Sustainable Behaviour

Awareness-behaviour link: Luque-Alcaraz et al. showed that nurses with higher environmental awareness are significantly more likely to engage in sustainable actions (p < 0.05). Barriers: Time constraints, insufficient resources (e.g., recycling bins), and limited leadership advocacy hinder translation of awareness into practice.

### 5. Data-Driven Staffing & Complexity Prediction

Bürgin et al. illustrated how routine administrative data can predict case complexity, informing needs-based staff planning. Although predictive accuracy is modest, calibrated models can support managers in aligning staffing levels with patient acuity, indirectly affecting nurse performance through workload balance.

**Table 2.**
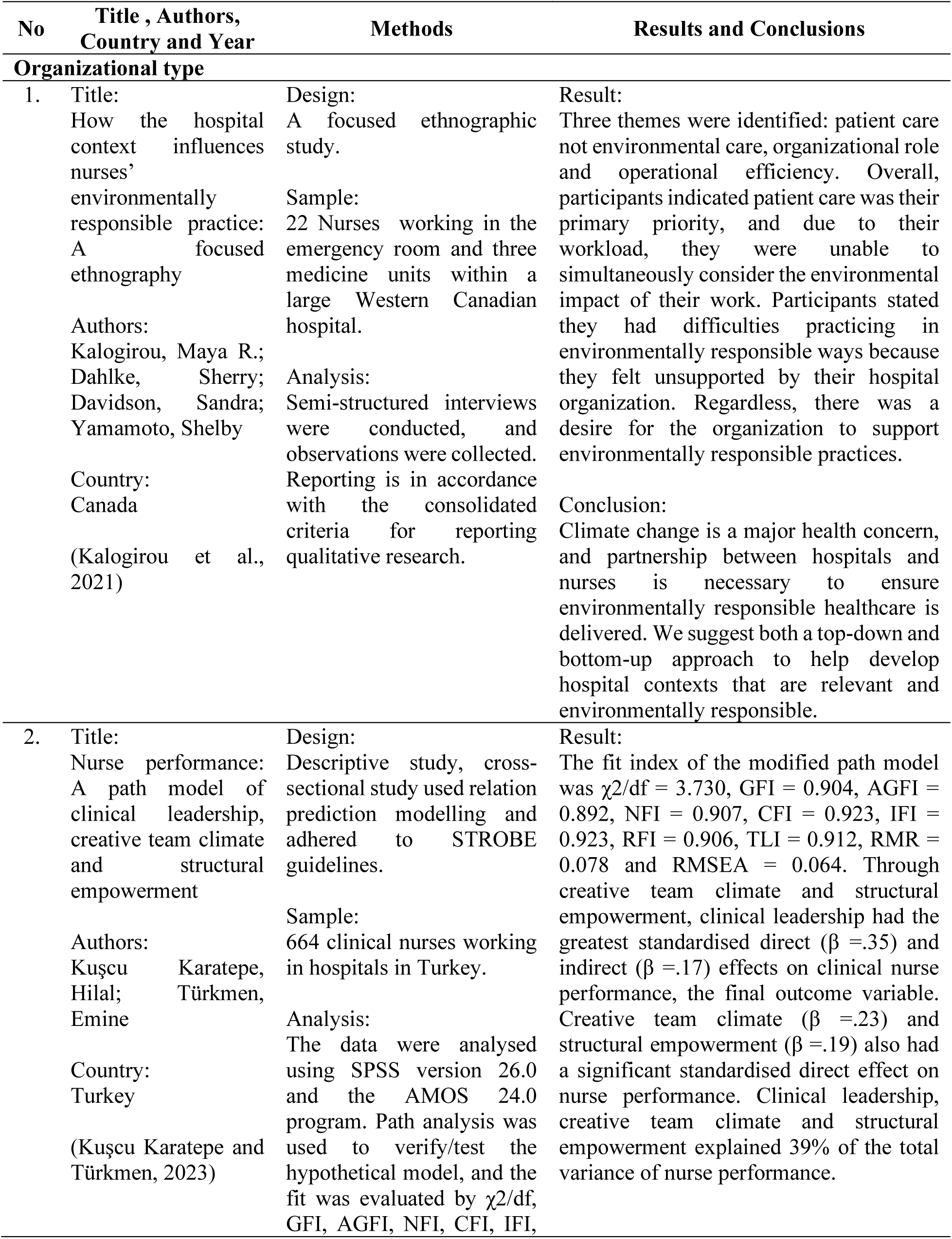
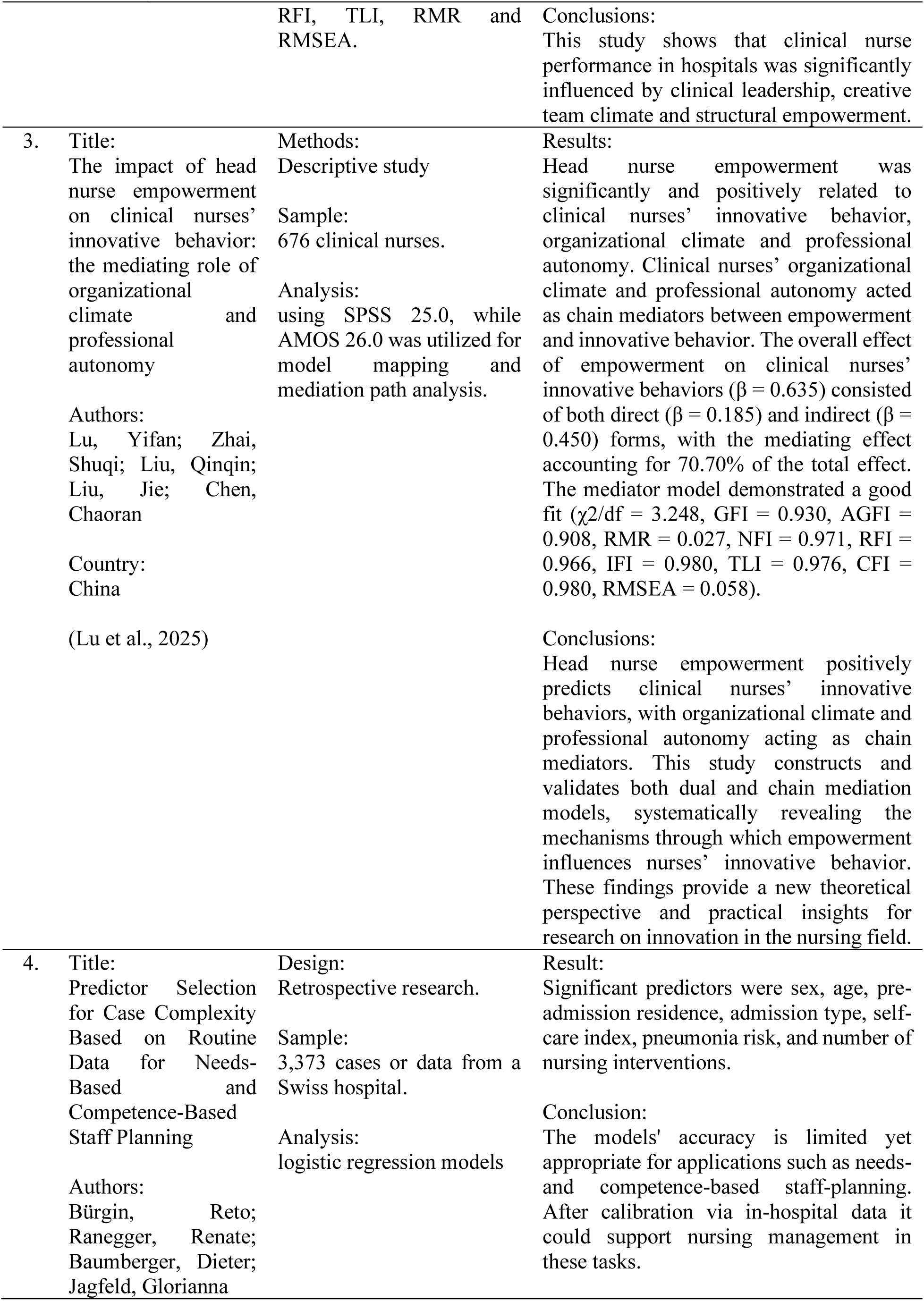
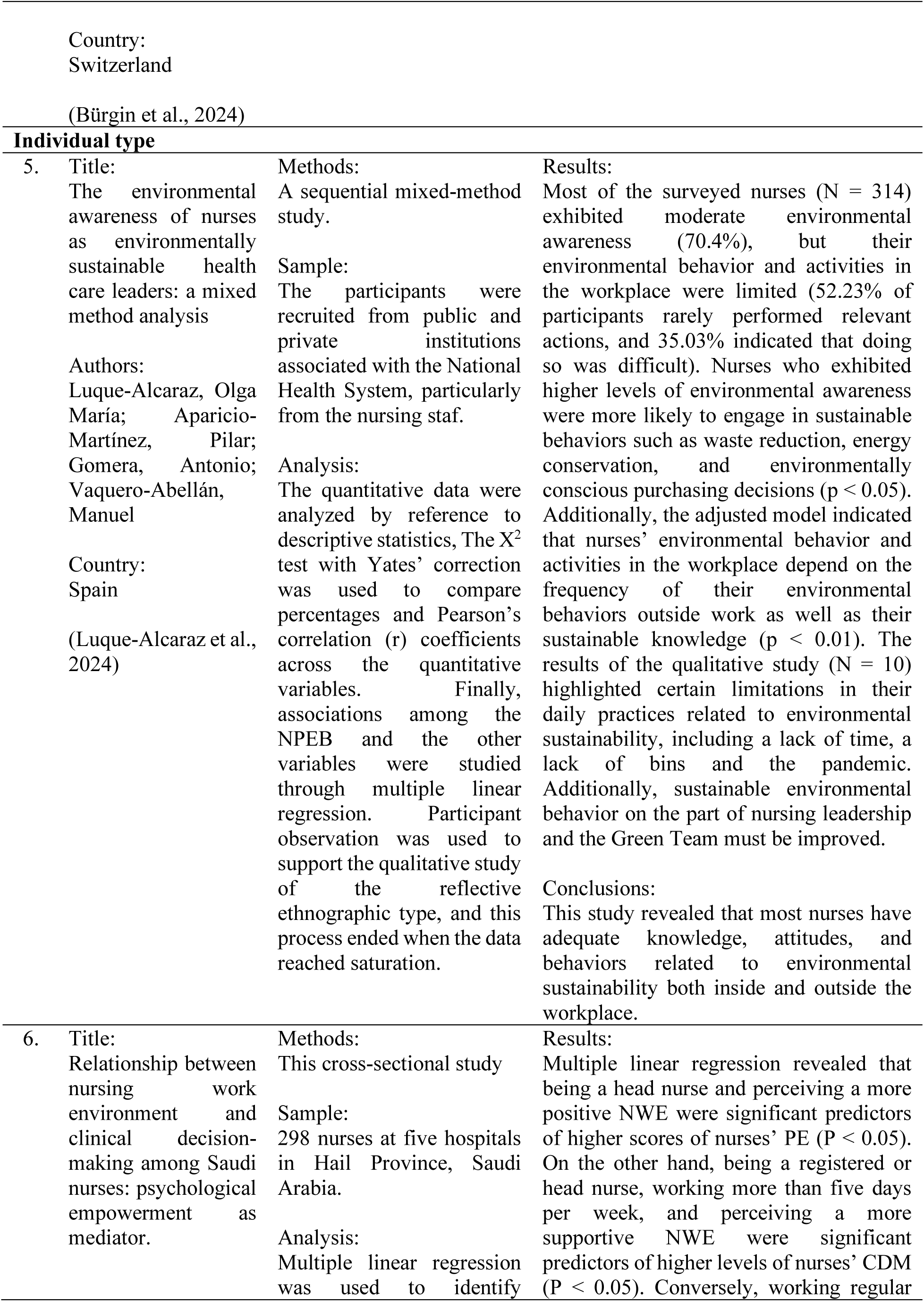
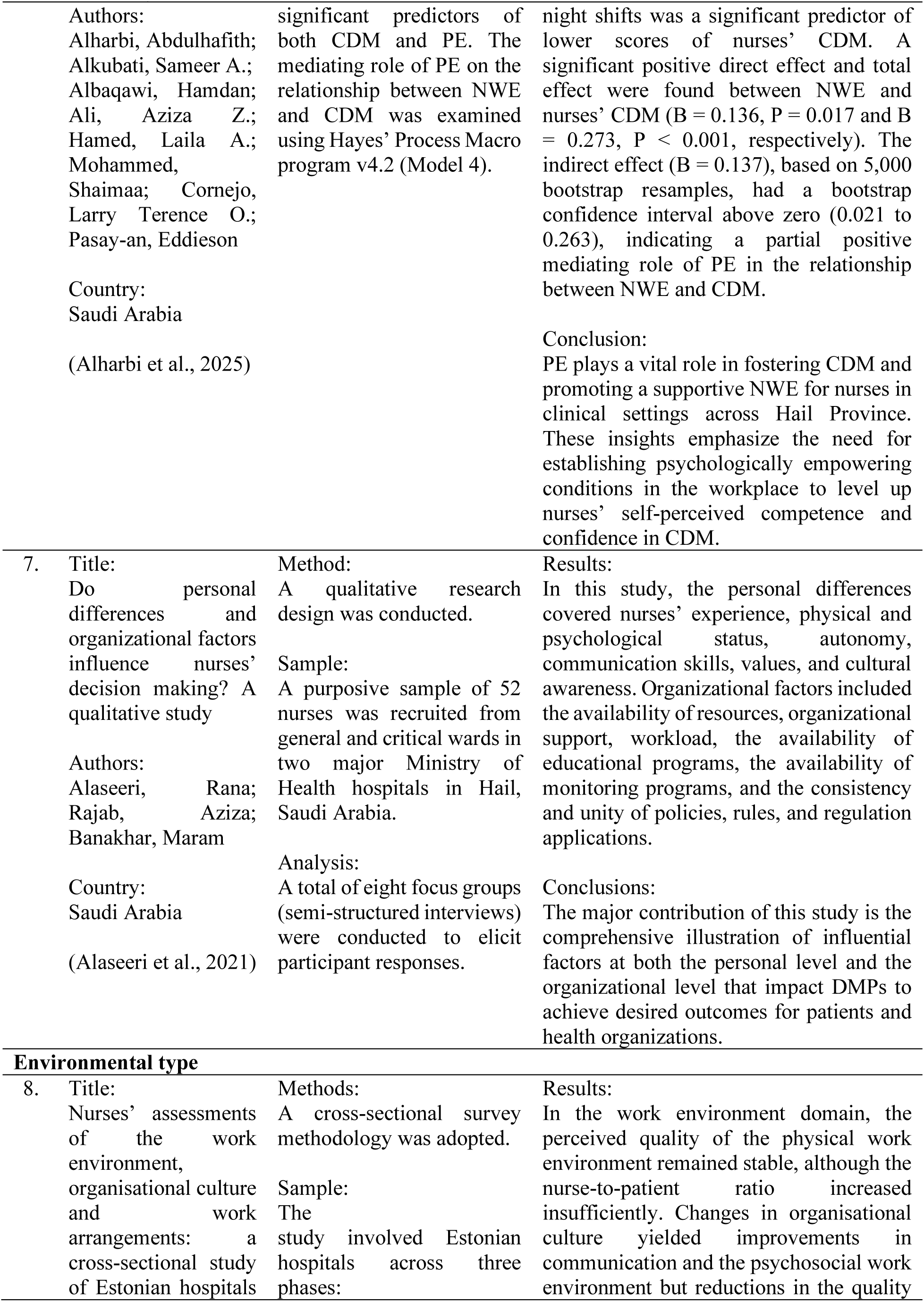
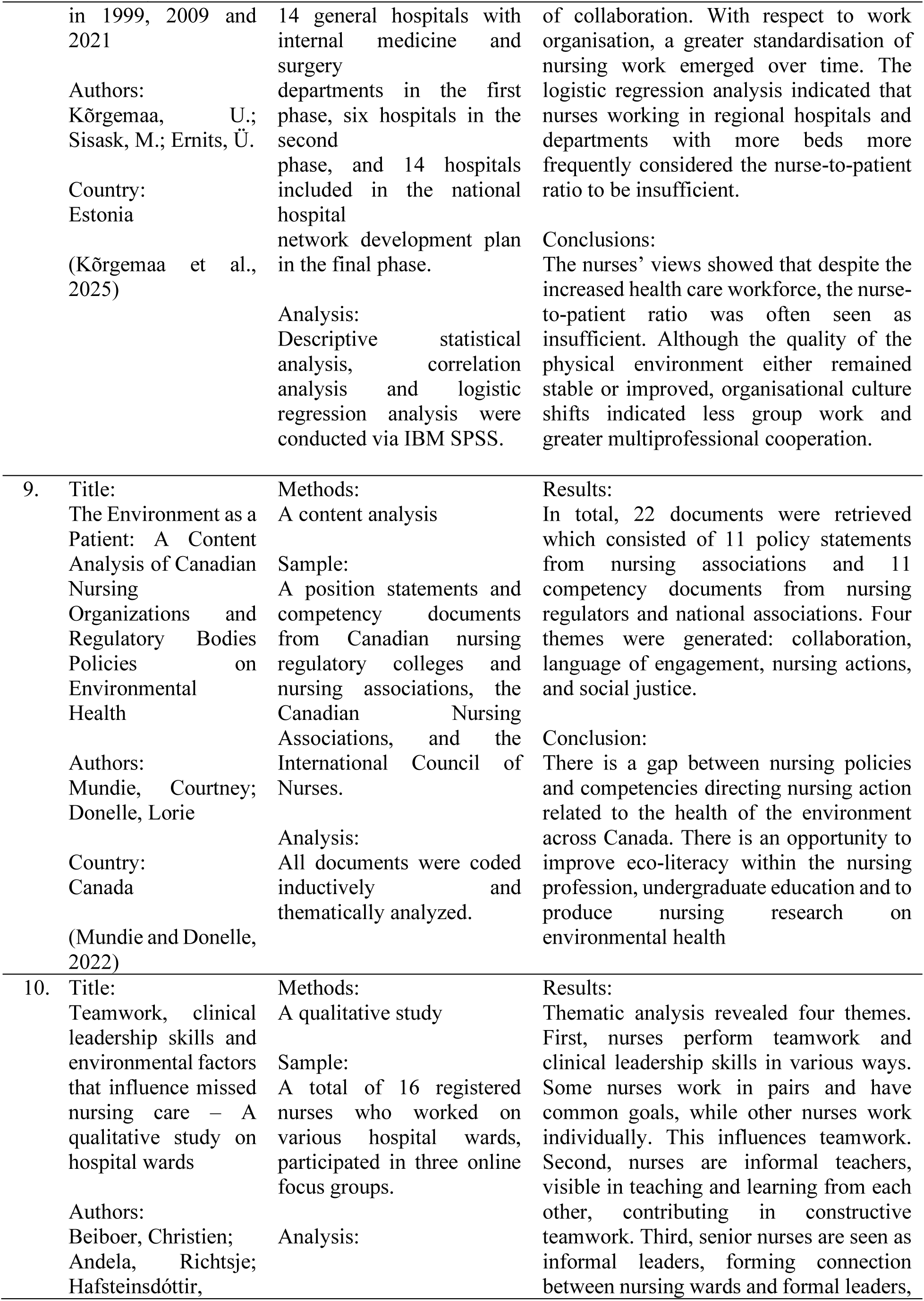
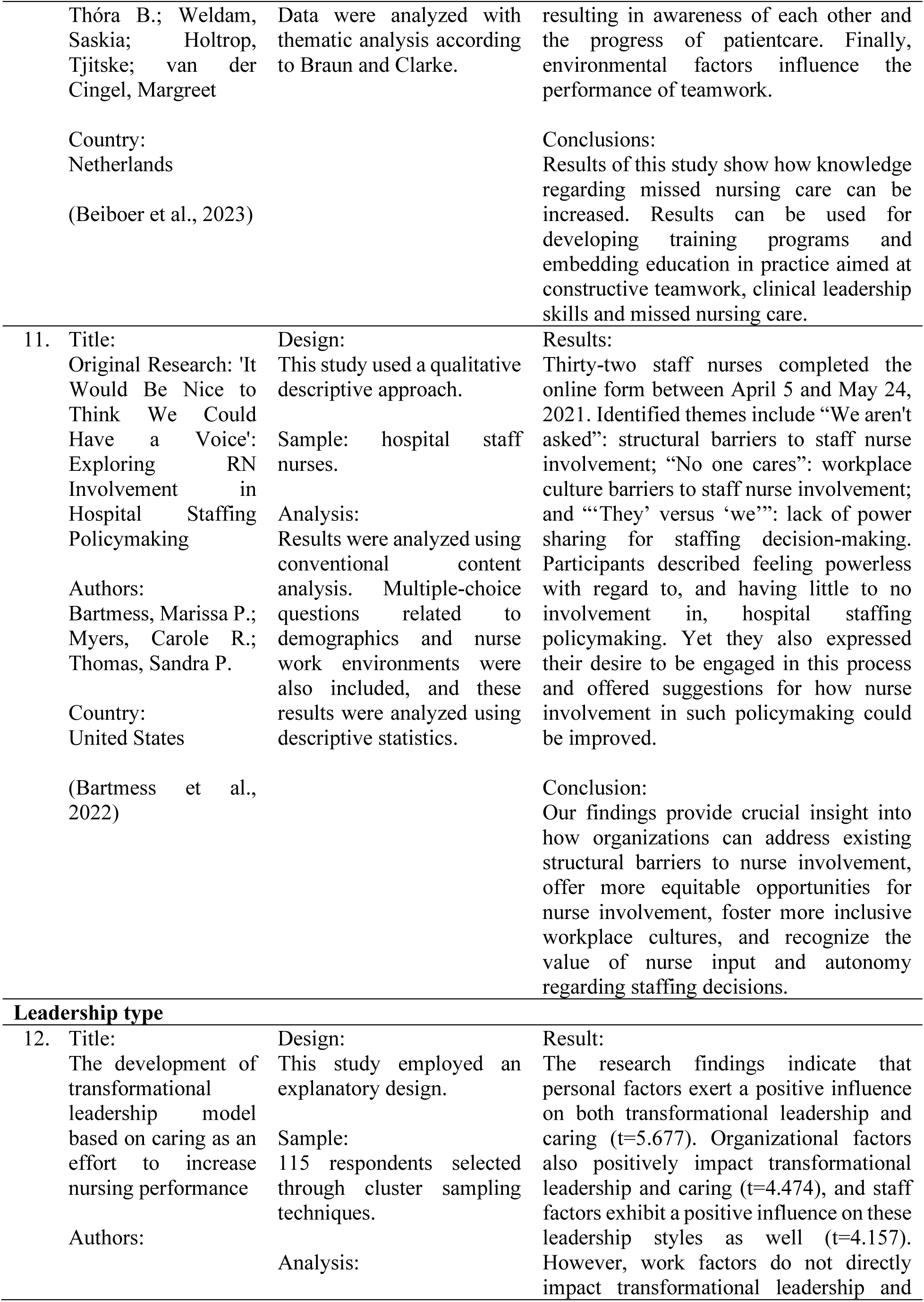
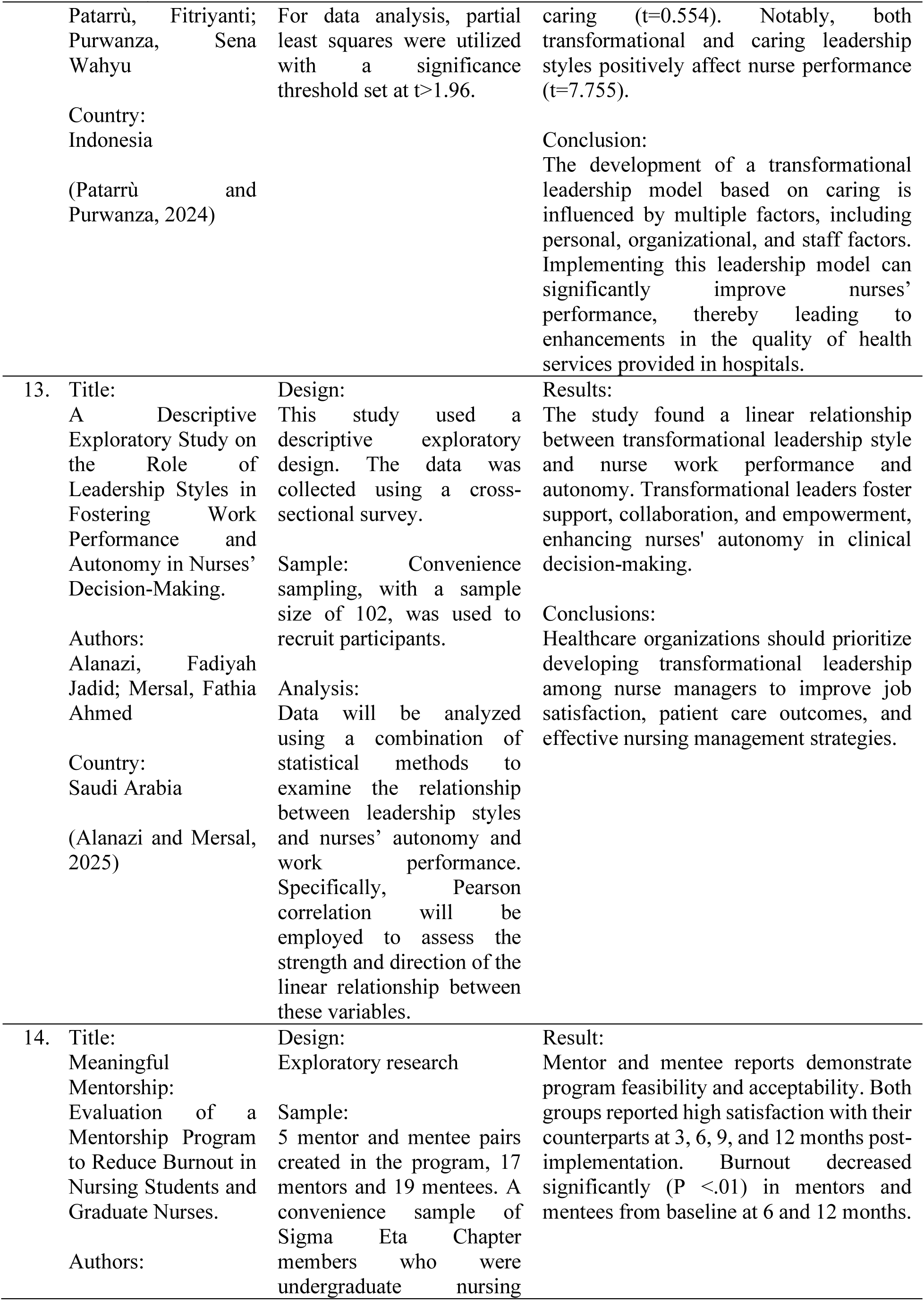
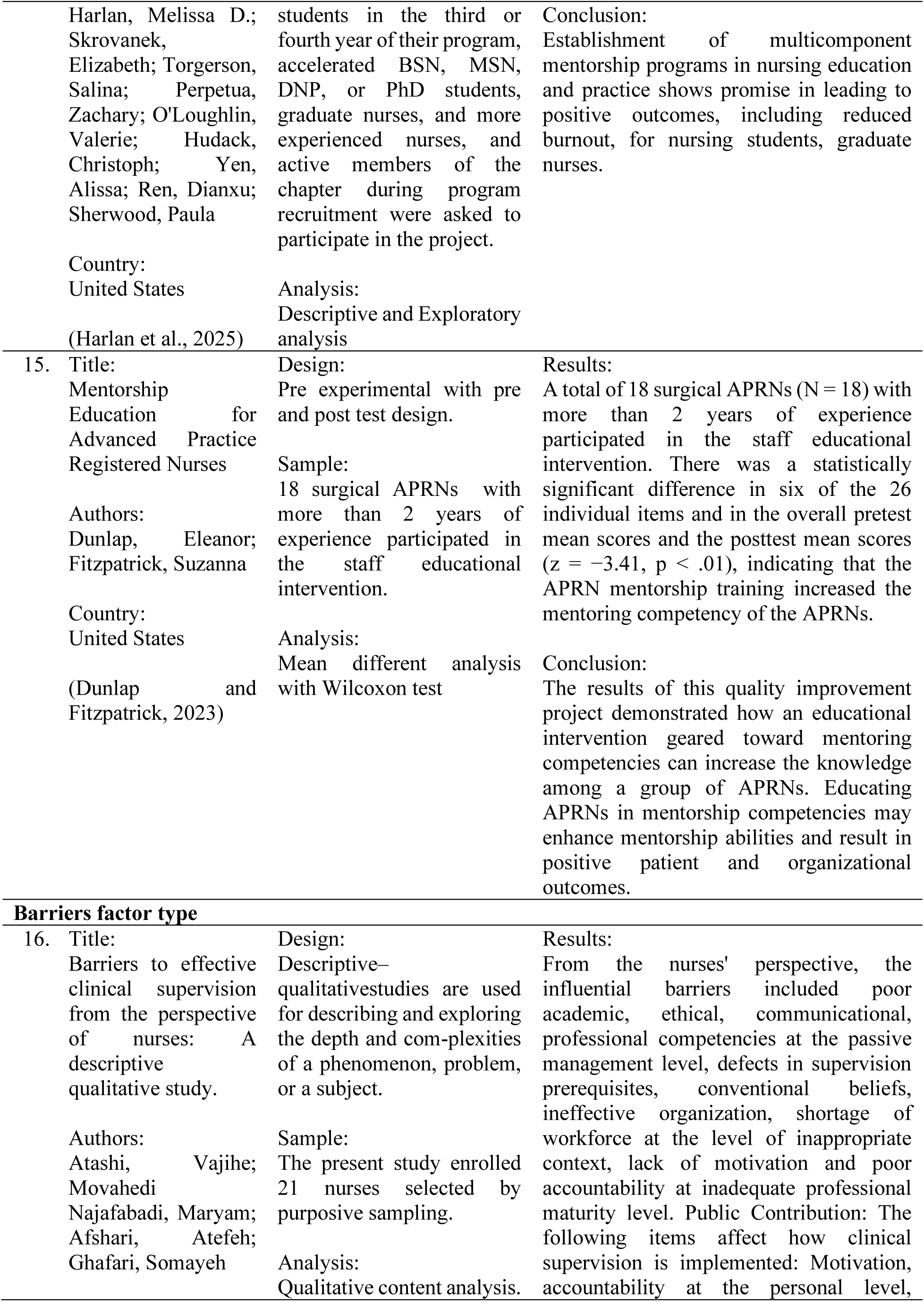
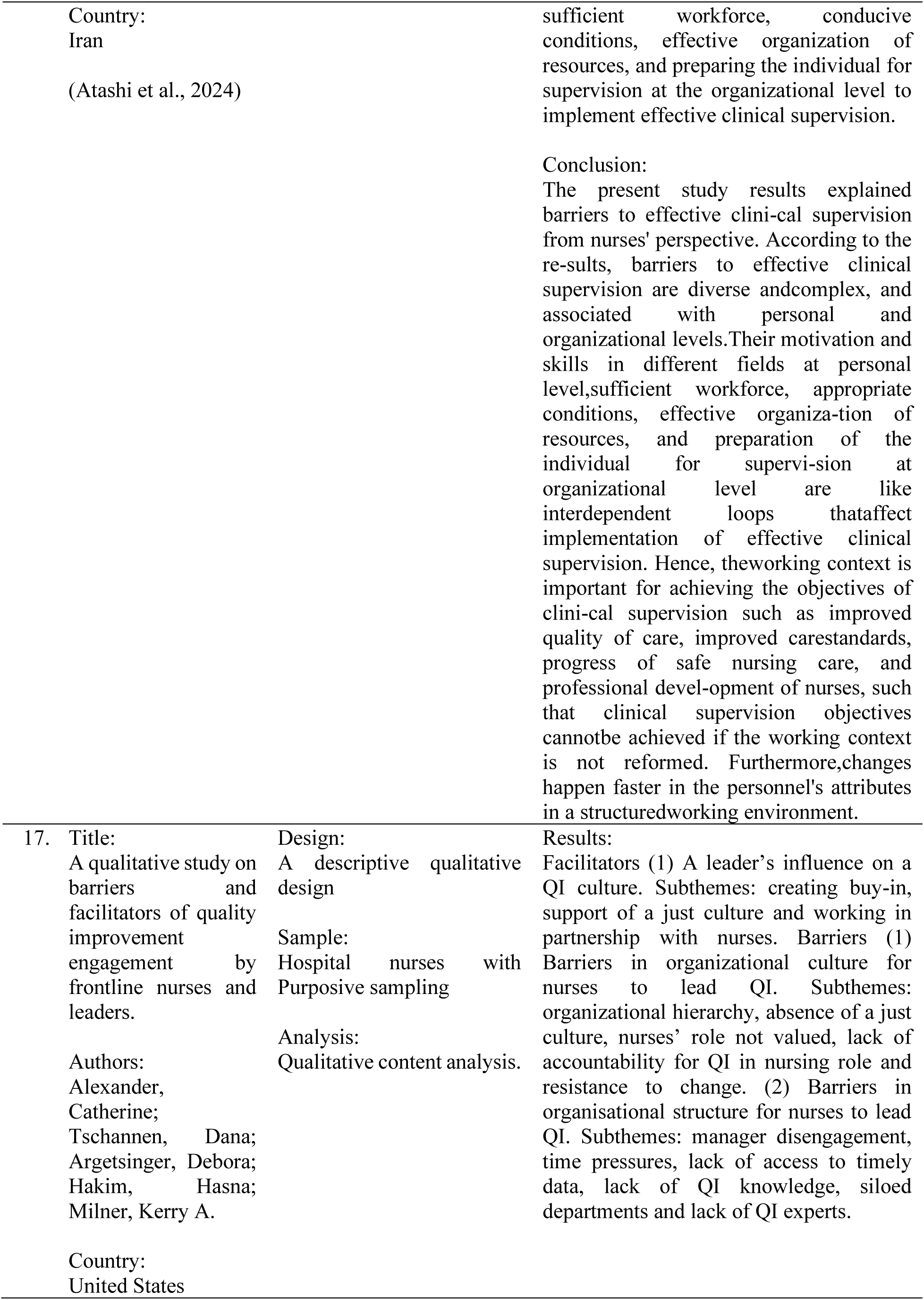
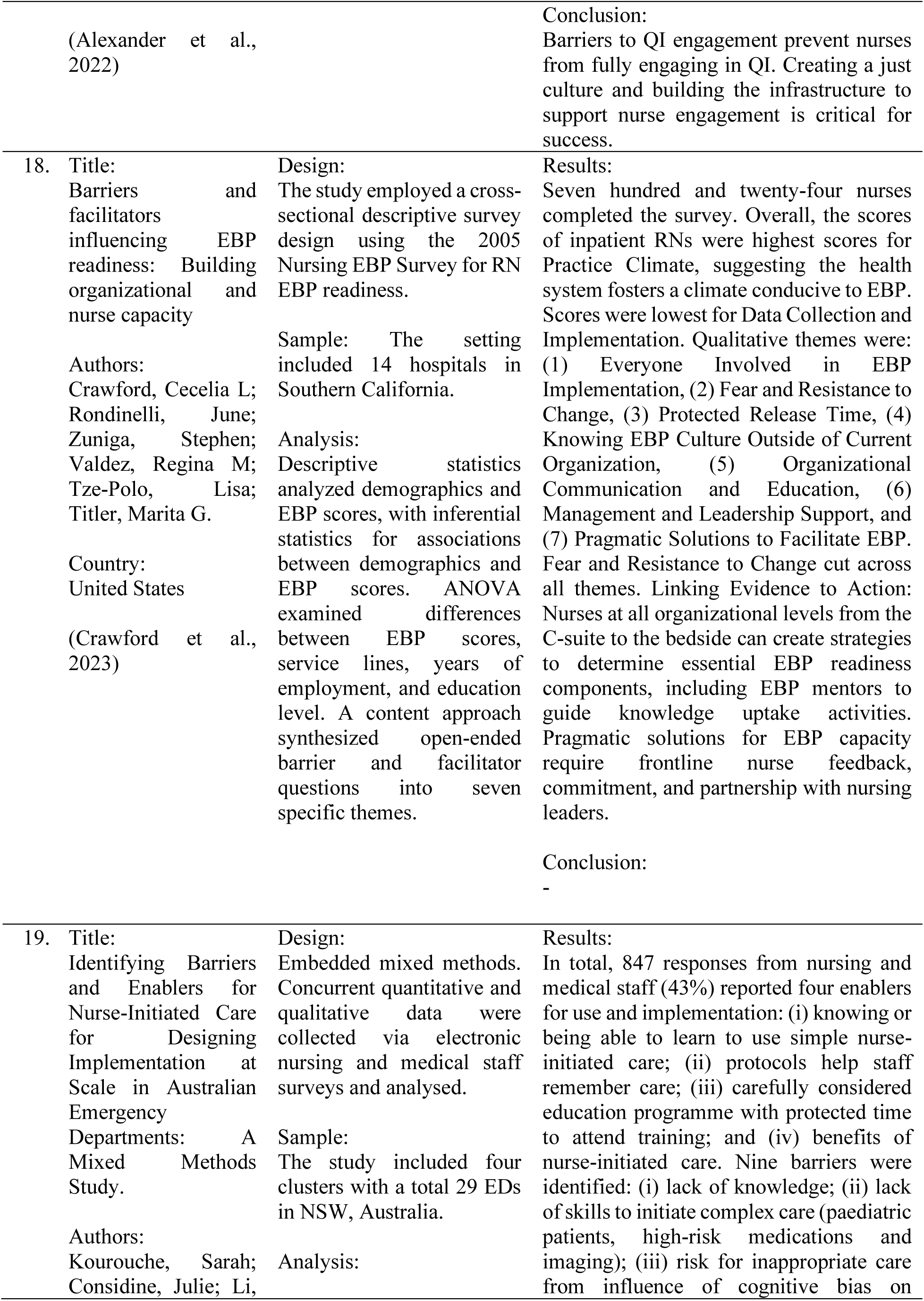
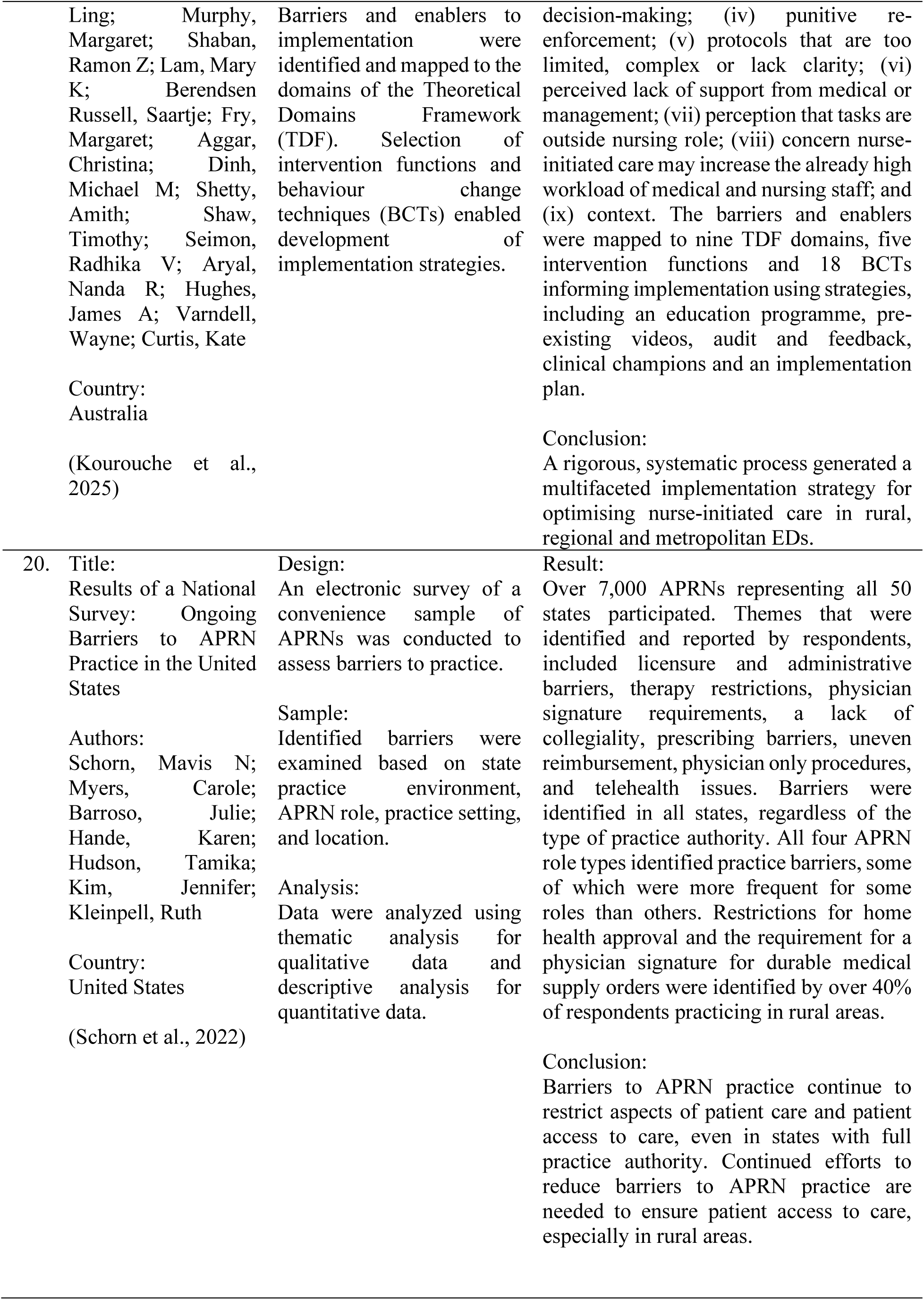
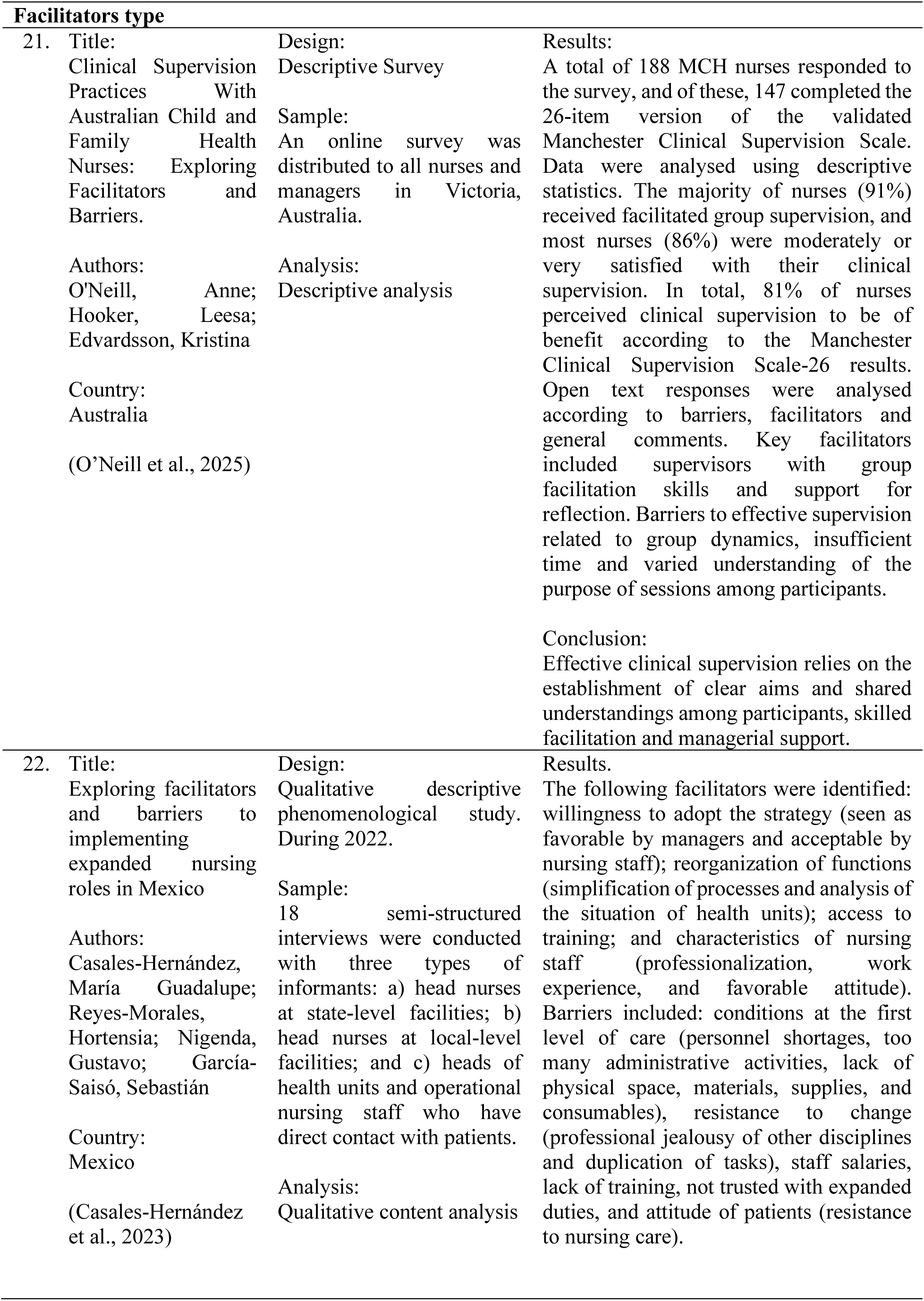
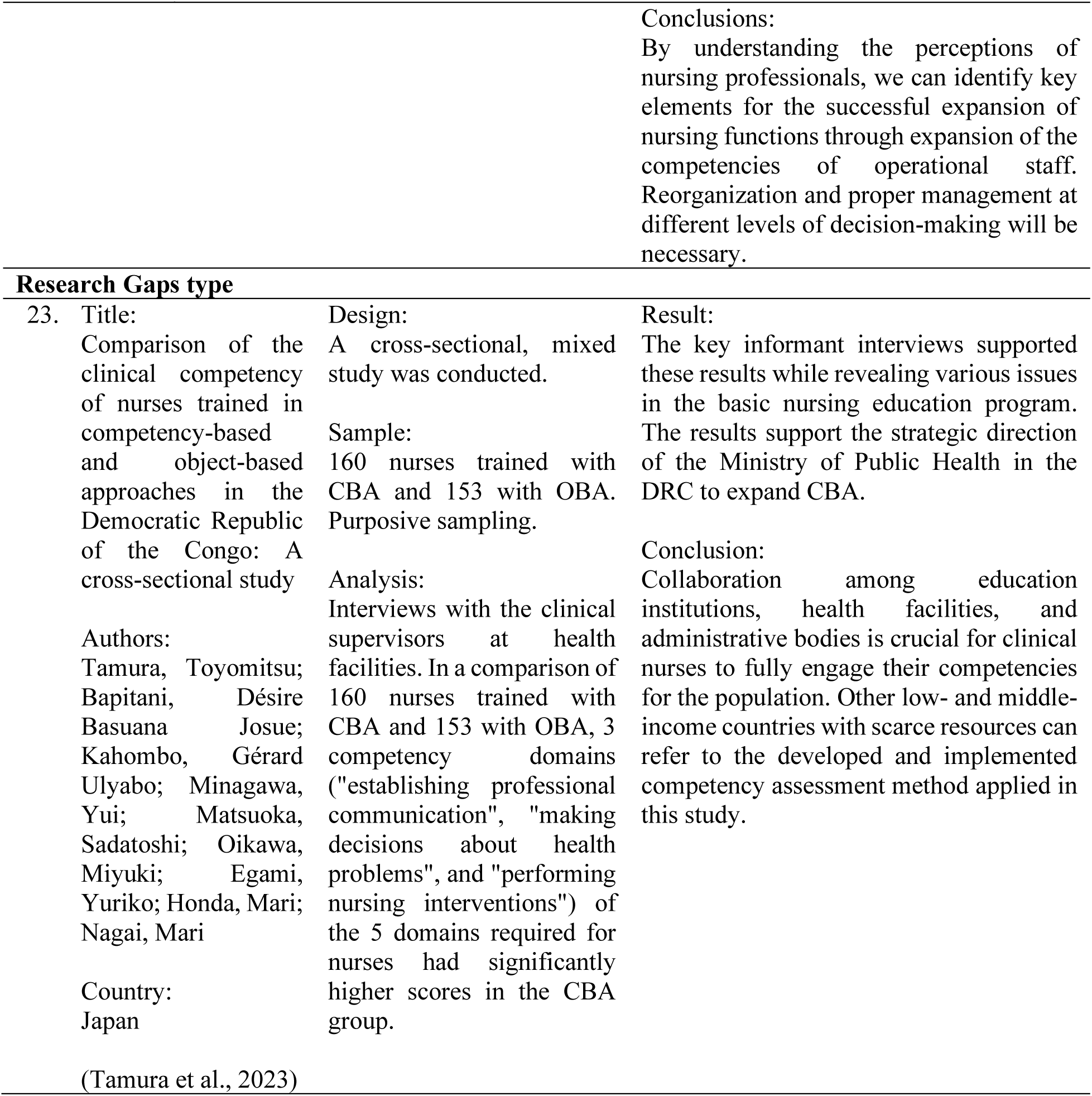
Results of Data Synthesis.

## Discussion

### 1. Clinical Leadership and Structural Empowerment

The empirical evidence consistently underscores clinical leadership as the most potent driver of nursing performance. Kuşcu Karatepe & Türkmen (2023) reported a standardized coefficient of β = 0.35, indicating that transformational leadership alone accounts for 39 % of the variance in nurse performance. Moreover, this effect is partially mediated through a creative team climate (β = 0.23) and structural empowerment (β = 0.19) (Kuşcu Karatepe and Türkmen, 2023).

## Conflict of Theory

Two dominant theoretical frameworks clash in interpreting these findings:

### 1.1. Transformational Leadership Theory (Bass, 1990)

Leaders inspire shared vision, foster intrinsic motivation, and empower followers to exceed routine expectations. Strongly supported: high β values, indirect pathways via empowerment and team climate.

### 1.2. Transactional Leadership Theory (Burns, 1978)

Leadership is based on contingent reward-punishment mechanisms that focus on extrinsic incentives. Weaker association (Lee et al., 2022) suggests limited impact on innovative nursing behaviors.

## Implications for Nursing Practice

### 1.1. Leadership Development Programs

Curricula for nurse managers should prioritize transformational competencies (vision-casting, coaching, individualized consideration) and include simulation-based training that demonstrates how to translate vision into concrete policies.

### 1.2. Organizational Architecture

Healthcare institutions must institutionalize mechanisms for structural empowerment transparent access to clinical data, participation in decision-making forums, and robust mentorship pathways.

### 1.3. Performance Measurement

Appraisal systems should incorporate validated instruments such as the Leadership Climate Survey and Kanter’s Empowerment Scale as predictive indicators of nursing performance alongside traditional clinical metrics.

### 2. Environmental Sustainability as a Dimension of Nursing Performance

The convergence of green nursing initiatives with traditional patient-care imperatives remains contested. Kalogirou (2021) identified a role-conflict where nurses prioritize direct patient care over environmentally sustainable actions, despite moderate environmental awareness. Conversely, Luque-Alcaraz (2024) highlighted the potential synergy that emerges when structural resources (e.g., automated waste-segregation systems) are provided.

## Conflict of Theory

### 2.1. Green Nursing Paradigm (McGain & Naylor, 2020)

Sustainability and quality care are complementary; nurses will adopt green practices when they recognize ecological benefits. Luque-Alcaraz (2024) demonstrate increased green behaviors when institutional support is present.

### 2.2. Task-Priority Model (Kahn et al., 1964)

High clinical workload monopolizes cognitive resources, diminishing capacity for additional tasks such as recycling. Kalogirou (2021) show reduced green actions in high acuity units.

## Implications for Nursing Practice

### 2.1. Policy Integration

Embed sustainability protocols directly into standard operating procedures (e.g., use of recyclable infusion sets, electronic documentation to reduce paper waste) so that green actions become part of routine care rather than an optional add-on.

### 2.2. Educational Enrichment

Revise undergraduate and continuing-education curricula to include Sustainability Science for Nurses, emphasizing evidence-based environmental interventions that do not compromise patient safety.

### 2.3. Outcome Metrics

Introduce sustainability indicators (e.g., kilograms of medical waste diverted, energy consumption per patient day) into the nursing performance dashboard, aligning incentives with environmental stewardship.

### 3. Innovation, Professional Autonomy, and Self-Determination

(Lu et al., 2025) demonstrated that head-nurse empowerment predicts innovative behavior (β = 0.635) with a mediation effect of 71 % through an empowering organizational climate and professional autonomy. This finding resonates with Self-Determination Theory (SDT) (Deci & Ryan, 2000), which posits that fulfillment of the basic psychological needs for autonomy, competence, and relatedness fuels intrinsic motivation and creative output.

## Conflict of Theory

### 3.1. SDT-Based Innovation Model

Autonomy and supportive climates directly stimulate innovation. Strong empirical support (high β, high mediation).

### 3.2. Resource-Constraint Model (Herzberg, 1966)

Innovation is inhibited when nurses face excessive workload and limited material resources. Evident in settings lacking adequate staffing or equipment, where empowerment does not translate into innovation.

## Implications for Nursing Practice

### 3.1. Empowerment Structures

Create formalized clinical practice councils where head nurses and frontline staff co-design workflow improvements, ensuring that decision-making authority is distributed.

### 3.2. Resource Allocation

Guarantee access to simulation labs, evidence-based practice tools, and time allowances for quality-improvement projects, mitigating the resource-constraint barrier.

### 3.3. Recognition Systems

Develop award programs that celebrate innovative nursing solutions, reinforcing the link between autonomy, competence, and professional identity.

### 4. Synthesis and Future Directions

The converging evidence delineates a triadic nexus:

### 4.1. Leadership that is transformational and structurally empowering

### 4.2. Sustainability integrated into clinical routines

### 4.3. Professional autonomy nurtured within a resource-rich, supportive climate

When these elements co-exist, they produce synergistic effects on nursing performance enhancing patient outcomes, fostering environmentally responsible practices, and catalyzing innovation. Conversely, the absence or misalignment of any component precipitates theoretical conflicts that manifest as performance deficits.

Future research should adopt multilevel longitudinal designs to examine how fluctuations in leadership style, empowerment policies, and sustainability initiatives jointly influence nurses’ intrinsic motivation and performance over time. Practically, health-care organizations are encouraged to holistically redesign their governance, operational, and educational frameworks to operationalize the integrated model presented herein.

## Conclusion

Recent literature (2021–2025) underscores that hospital nurses’ performance within their clinical authority is shaped by a dynamic interplay of organizational, individual, and environmental factors. Key determinants include supportive leadership, manageable workloads, empowerment, and a positive organizational climate. Barriers such as hierarchical culture, resource shortages, and lack of support persist, but can be mitigated through comprehensive leadership development, mentorship, and culture change interventions. There is an urgent need for validated, context-sensitive measurement tools and research in diverse settings, particularly LMICs, to optimize nurse performance and clinical authority.

## Data Availability

All data produced in the present work are contained in the manuscript

## Notes

### Competing Interest Statement

The authors have declared no competing interest.

### Funding Statement

This study did not receive any funding

